# A Randomized Clinical Trial of Metformin to Reduce Frailty in Older Adults with Glucose Intolerance

**DOI:** 10.64898/2026.07.27.26359055

**Authors:** Nicolas Musi, Chen-Pin Wang, Daniel MacCarthy, Zhaohong Feng, Jacob T. Holmes, Masayoshi Suda, Tamar Pirtskhalava, Arianne Aslamy, Jonathan Wanagat, Robert Brooke, Tamar Tchkonia, James L. Kirkland, Steve Horvath, Sara E. Espinoza

## Abstract

**Importance:** Preclinical and human observational studies suggest that metformin may decrease age-related pathology, including frailty.

**Objective:** Determine whether metformin reduces frailty progression and biological age in older adults with glucose intolerance, a population at increased risk of becoming frail.

**Design, Setting and Participants:** Randomized, double-blind, placebo-controlled trial of metformin in 145 non-frail or pre-frail older adults. Participants (72±5 years, 48% female, 94% White, 35% Hispanic) were randomized to metformin *vs*. placebo for two years.

**Main Outcomes and Measures:** Effect on frailty was primarily determined using generalized estimating equations by change in the Fried frailty phenotype score (based on weight loss, exhaustion, physical activity, gait speed, and grip strength). Because metformin can cause significant weight loss, effects on the Fried score were assessed with and without the weight loss criterion. Frailty also was assessed by change in the frailty index (composite of 95 deficits). Biological age was estimated by DNA methylation-based epigenetic clocks in blood.

**Results:** Metformin led to a non-linear response in the Fried score rate of change, with an upward trajectory in year 1 (0.72±0.22 per year vs. placebo, p=0.0011) and stabilization in year 2 (−0.33±0.17 per year vs. placebo, p=0.056). Metformin led to more weight loss than placebo (−5.7±5.2 *vs*. −2.3±5.4 kg, p=0.0002); thus, when assessing effect on Fried score without the weight loss criterion, no difference was observed, indicating that weight loss in year 1 accounted for the change in Fried score. Notably, metformin caused a steady improvement in the frailty index (−0.006±0.0026 per year vs. placebo, p=0.0222) that persisted with covariates adjustment including body mass index. Metformin reduced biological age estimated by PC-Horvath2 (−0.40±0.16 per year, p=0.014) and PC-Hannum (−0.33±0.16 per year, p=0.047) clocks. Metformin was well tolerated.

**Conclusions and Relevance:** Metformin halts the progression of the deficit accumulation frailty index and reduces biological age, suggesting potential benefit for extending healthspan.

**Trial Registration:** ClinicalTrials.gov: NCT02570672.

**Key Points:** *Question:* Does metformin slow progression of frailty and biological aging in older adults?

*Findings:* In this randomized, placebo-controlled clinical trial of 145 non-frail or pre-frail older adults with glucose intolerance (prediabetes), metformin led to a significant reduction in the deficit accumulation frailty index but not the Fried frailty phenotype. Metformin also reduced biological age estimated by the epigenetic clocks PC-Horvath2 and PC-Hannum in peripheral blood.

*Meaning:* Metformin treatment in older adults improves frailty index and indicators of biological age, suggesting potential benefits for extending healthspan.

## INTRODUCTION

Frailty is a syndrome of physical decline and vulnerability that affects a high proportion of older adults and increases risk of morbidity and disability, leading to increased health care costs.^1,2^ Glucose intolerance (prediabetes) and diabetes, highly prevalent in older adults,^3^ are among the strongest frailty risk factors.^4,5^

Metformin has emerged as a promising intervention to extend years of life spent free of disease, disability, and frailty (*i.e*. healthspan).^6–8^ In addition to improving glucose homeostasis, metformin has pleiotropic effects on several hallmarks of aging, including epigenetics, cellular senescence, and inflammation,^9–11^ which are implicated in the pathophysiology of age-related diseases and frailty.^12^ Preclinical and observational studies suggest metformin reduces age-related disease, mortality, and frailty.^9,13–21^ Metformin is a particularly attractive gerotherapeutic because it has been in clinical use for decades, has excellent safety and tolerability profiles, and is low cost. However, to date, no long-term clinical trial has been conducted to examine effects of metformin on aging-focused clinical outcomes or markers of biological aging.

Here, we report results from a randomized, placebo-controlled clinical trial to determine whether metformin slows frailty progression in older adults with glucose intolerance, a population at high risk of becoming frail. We also examined the effects of metformin on DNA methylation-based biological aging clocks.

## METHODS

### Overview

This was a single-site, randomized (1:1 allocation), double-blind, placebo-controlled, parallel-group trial, conducted from April 2016 to January 2024 at the Audie L. Murphy Veteran Affairs (VA) Hospital, in San Antonio, TX, USA. The trial was registered with clinicaltrials.gov (NCT02570672) and approved by the local Institutional Review Board and an NIH-appointed Data and Safety Monitoring Board. All participants provided written informed consent. The study was conducted in accordance with Good Clinical Practice and the Declaration of Helsinki. The study design has been previously described.^22^

### Participant eligibility

We studied non-frail or pre-frail older adults, based on the Fried frailty phenotype,^23,24^ who were glucose intolerant. Potential participants were recruited by several methods, including flyers, ads, and referrals. Interested individuals underwent telephone screening and those without clear exclusionary criteria were invited for in-person screening at which a medical history, physical examination, and a 75 g oral glucose tolerance test (OGTT) were performed. Glucose intolerance was defined based on a 2-hour glucose concentration of 140-199 mg/dL. Participants were assessed for cognitive impairment and depressive symptoms with the Mini Mental State Examination (MMSE) and Geriatric Depression Scale (GDS), respectively. The Fried frailty phenotype was assessed by 10-foot walk testing, grip strength, the Minnesota Leisure Time Physical Activity Questionnaire, the GDS for exhaustion, and unintentional weight loss of ≥10 pounds in the prior year (Table S1). Individuals were characterized as frail if they showed ≥3 or more of: 1) slow gait speed, 2) muscle weakness, 3) low physical activity, 4) exhaustion, and 5) unintentional weight loss as described.^23,24^ Blood and urine samples were analyzed for complete blood count, comprehensive metabolic panel with liver function, hemoglobin A1c, vitamin B12, and urinalysis, by the clinical laboratory at Audie L. Murphy VA Hospital. Participants were excluded if they were not glucose intolerant, were frail, or if they had glomerular filtration rate (eGFR) <45 mL/min (Table S2).

### Randomization

Participants meeting eligibility criteria were randomized in a design balanced by sex and ethnic group (Hispanic *vs*. Non-Hispanic White or any other group) using a random number generator by a research pharmacist who was the only person with the randomization information.

### Intervention and study procedures

The intervention was 24 months of oral metformin (immediate release) or an indistinguishable placebo, initiated at 500 mg per day and increased by 500 mg approximately every 2 weeks to a maximum tolerated dose of up to 2,000 mg/day. Participants who required dose reduction due to gastrointestinal symptoms were re-challenged to the maximum tolerated dose. Before study drug initiation, all participants received diet and exercise counseling^25,26^ from a registered nurse and certified diabetes care and education specialist. All visits were in person except during the COVID-19 pandemic (March 2020–June 2020), when in-person visits were paused and visits were conducted by telephone and study drug mailed to participants’ homes. Because dosing fluctuated based on tolerability, we calculated the average daily dose according to pill counts (or phone reports during the COVID-19 pandemic).

After study initiation, medical histories and physical examinations, frailty assessments, and OGTTs were repeated every 6 months. Safety laboratory (complete blood count, comprehensive metabolic panel with renal and liver function) and hemoglobin A1c were measured every 3 months. Serum vitamin B12 was measured every 12 months. Serum concentrations of growth differentiation factor (GDF)15 were measured every 6 months, as described.^27^

### Safety considerations

If eGFR was <45 mL/min at any point, the study drug was withheld and eGFR was rechecked in two weeks. The study drug was resumed at the prior dose if the repeat eGFR was >45 mL/min, initiated at a reduced dosage of maximum of 1,000 mg/day if the repeat eGFR was 30-45 mL/min, or discontinued permanently if the repeat eGFR was <30 mL/min.

### Frailty assessments

The primary outcome of frailty was assessed by the rate of change in the Fried phenotype score, which ranges from 0 to 5, with a higher score indicating greater frailty. Considering the known weight-lowering effect of metformin, post-randomization Fried phenotype was assessed at baseline for all criteria, except weight loss was defined as loss of ≥5% of initial body weight during follow-up. We also assessed frailty by the deficit accumulation frailty index (composite of 95 deficits), calculated based on assessment of chronic diseases, conditions, impairments, and disability using validated criteria (Table S3).^28,29^ The frailty index is the sum of all deficits divided by the number of items assessed, and ranges from 0 to 1, with a higher score indicating greater frailty.

### Diabetes conversion ascertainment

If any of the following occurred during the study, a 2-hour OGTT was performed to confirm diabetes conversion: 1) fasting glucose ≥126 mg/dL, 2) hemoglobin A1c ≥6.5%, 3) glucose ≥200 mg/dL on 2-hour OGTT, or 4) clinical signs and symptoms of diabetes. Conversion to diabetes was ascertained if the 2-hour glucose was ≥200 mg/dL on the repeat OGTT. An independent physician then initiated open-label metformin treatment at 500 mg daily titrated to a maximum tolerated dose up to 1,000 mg twice daily. All participants and study staff except the study pharmacist remained blinded to drug assignment.

### DNA methylation and epigenetic clock assessments

DNA was isolated from EDTA collected buffy coat using QIAamp DNA kit. DNA methylation was assessed using Infinium MethylationEPIC BeadChip arrays (Illumina, San Diego, CA). DNA was bisulfite converted (Zymo EZ DNA methylation kit), processed according to the manufacturer’s protocol, and scanned for methylation readout as described.^30^ Raw methylation data were imported using the *minfi* (v1.40.0) R package, corrected for technical variation, and normalized using the Noob method. Conventional and principal component (PC)-based epigenetic clocks, which have been found useful for personalized medicine and longitudinal tracking of aging processes,^31^ were calculated from normalized data. Clocks examined included PC-Horvath2, PC-PhenoAge, Hannum, PC-Hannum, Robust Horvath Pan-Tissue, PC-GrimAge, Levine PhenoAge, GrimAge (real-age variant), PC-DNAmTL, DNAmTL, DunedinPACE, GrimAge v2, and DNAmFitAge.^32,33^

### Power analysis and sample size

Sample size was estimated based on preliminary data derived from a prior observational cohort study^4^ indicating that the Fried score worsened in 53% of participants over 18 months. Therefore, *a priori* power analysis with significance level of α=0.05 indicated that if 120 participants completed the trial, we would have 86% power to detect a meaningful difference in Fried score in the metformin *vs*. placebo groups.^22^ Overall effect size was projected by incorporating the possibility of noncompliance and treatment arm switching due to diabetes conversion.

### Statistical analysis

Between-group differences in change of body weight, body composition, physical function, and metabolic indices from baseline to 12 and 24 months were analyzed using Wilcoxon rank sum tests. Frailty progression was defined as worsening of Fried phenotype score and frailty index. Primary inferences were drawn based on intention-to-treat analyses, including randomized participants who received at least one dose of study drug. The effect of metformin on frailty progression was assessed with generalized estimating equations (GEE) models with terms for time, treatment group, and their interaction. Models were examined unadjusted and adjusted for baseline and time-varying covariates, including age, sex, race, ethnic group, cholesterol, triglycerides, fasting glucose, and body mass index (BMI). The optimal temporal trend and working correlation structure were selected using the quasi-likelihood under the independence model criterion (QIC). The final GEE models used the temporal trend and working correlation structure with the lowest QIC for each frailty outcome.

Missing data were minimal; primary analyses used observed data. In view of the well-known effect of metformin to reduce body weight, change in Fried phenotype was also evaluated without the weight loss criterion. Because the study included prediabetic individuals, sensitivity analyses were done with censored observations after diabetes conversion among those who converted to diabetes.

Robust epigenetic clock versions were calculated as described^31^ to reduce technical noise and improve reliability for longitudinal tracking. Outcomes were analyzed using linear mixed-effects models: Epigenetic Age ∼ Time × Treatment + Chronological Age + Sex + (1|Subject), with treatment × time interaction as the primary coefficient of interest. Mixed-effects models were used to account for within-subject correlation from repeated measurements and subject-specific baseline heterogeneity, while preserving inference on the average treatment effect over time.

All analyses were carried out using SAS 9.4, except for analyses of epigenetic clocks, which were carried out with R *minfi* v1.40.0 for methylation processing; mixed-effects models via lme4/nlme).

## RESULTS

### Participants

After pre-screening 1,500 individuals, 388 underwent in-person screening for eligibility (Figure 1), 149 participants met criteria for study entry, and 145 were randomized (72 metformin, 73 placebo). Four participants (2 metformin, 2 placebo) declined participation after randomization and did not take study medication. Therefore, 141 participants (70 metformin, 71 placebo) were included in the intention-to-treat analysis. Participants were 48% female, 94% White, 35% Hispanic/Latinx, 4.3% African American, and 2.1% Other, with mean age 72 ±5 years and BMI 30.9 ±5.9 kg/m^2^ (Table 1).

**Figure 1.**
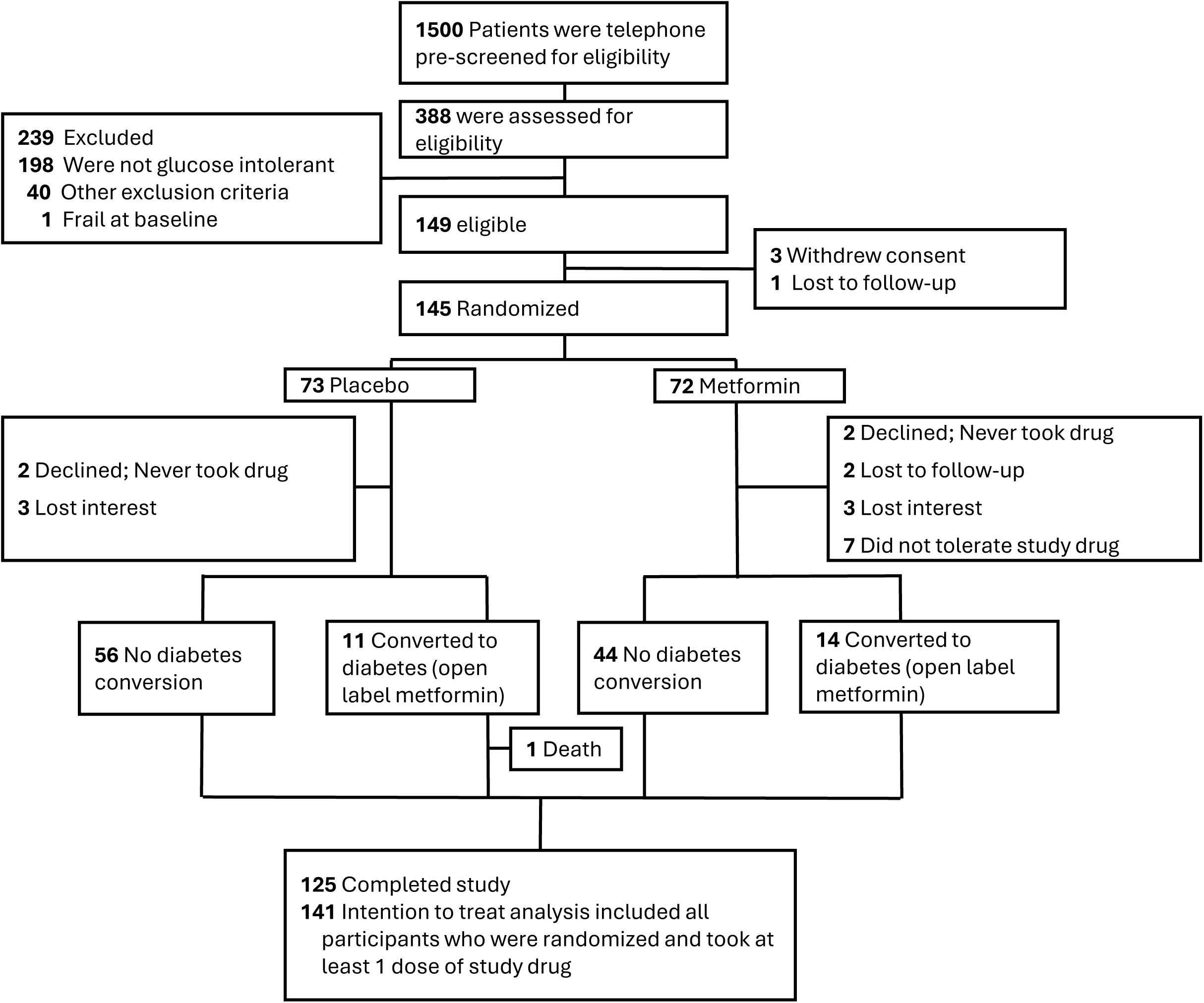
CONSORT diagram

**Table 1.** Demographic and clinical characteristics of participants at baseline.

|  |  |  |  |
| --- | --- | --- | --- |
|  | Metformin | Placebo | Overall |
|  | N= 70 | N= 71 | N= 141 |
| Characteristic | Mean (SD) or N (%) |  |  |
| Age, years | 71.0 (5.4) | 72.6 (5.1) | 71.8 (5.3) |
| **Sex** |  |  |  |
| Women | 34 (49) | 34 (48) | 68 (48) |
| Men | 36 (51) | 37 (52) | 73 (52) |
| **Race** |  |  |  |
| White | 64 (91) | 68 (96) | 132 (94) |
| African American | 3 (4.3) | 3 (4.2) | 6 (4.3) |
| Other | 3 (4.3) | -- | 3 (2.1) |
| **Ethnicity** |  |  |  |
| Hispanic/Latinx | 27 (39) | 22 (31) | 49 (35) |
| Not Hispanic/Latinx | 43 (61) | 49 (69) | 92 (65) |
| **Physical assessment, clinical laboratory** |  |  |  |
| Body weight, kg | 88.7 (19.8) | 85.7 (18.1) | 87.2 (18.9) |
| Body mass index, kg/m<sup>2</sup> | 31.1 (5.5) | 30.7 (6.3) | 30.9 (5.9) |
| Waist circumference, cm | 106.4 (17.3) | 102.3 (13.6) | 104.3 (15.6) |
| Systolic blood pressure, mmHg | 136.7 (15.4) | 139.4 (13.8) | 138.1 (14.6) |
| Diastolic blood pressure, mmHg | 73.6 (8.8) | 75.2 (9.6) | 74.4 (9.2) |
| Fasting glucose, mg/dL | 100.7 (9.2) | 100.0 (10.5) | 100.4 (9.9) |
| 2-hour glucose on oral glucose tolerance | 166.5 (17.6) | 169.5 (17.4) | 168.0 (17.5) |
| test, mg/dL |  |  |  |
| Hemoglobin A1c, % | 5.7 (0.4) | 5.7 (0.4) | 5.7 (0.4) |
| Total cholesterol, mg/dL | 170.2 (46.5) | 167.8 (38.0) | 168.9 (42.3) |
| High density lipoprotein, mg/dL | 50.2 (13.5) | 55.2 (17.6) | 52.7 (15.8) |
| Low density lipoprotein, mg/dL | 95.0 (35.6) | 91.0 (31.8) | 93.0 (33.7) |
| Triglycerides, mg/dL | 130.0 (61.1) | 110.0 (43.9) | 119.9 (53.9) |
| Glomerular filtration rate, mL/min/1.73 m<sup>2</sup> | 66.6 (13.4) | 64.7 (10.8) | 65.6 (12.1) |
| Aspartate aminotransferase, IU/L | 21.1 (6.3) | 20.9 (6.5) | 21.0 (6.4) |
| Alanine aminotransferase, IU/L | 21.2 (8.8) | 20.1 (10.2) | 20.6 (9.5) |
| Mini Mental State Exam, score | 28.6 (1.3) | 28.9 (1.2) | 28.7 (1.3) |
| Geriatric Depression Scale, score | 1.24 (1.6) | 1.28 (1.5) | 1.26 (1.5) |
| Short Physical Performance Battery, score | 10.1 (1.3) | 10.3 (1.2) | 10.0 (1.3) |
| Gait speed, meter/sec, fastest 10-foot walk | 1.16 (0.20) | 1.14 (0.17) | 1.15 (0.18) |
| Frailty Index | 0.17 (0.04) | 0.17 (0.04) | 0.17 (0.04) |
| Fried Phenotype, score | 0.39 (0.60) | 0.45 (0.60) | 0.42 (0.60) |

### Follow-up

The median follow-up time was 2.08 years (mean=1.95 ±0.73 years). Despite the blinded design, and in line with previous studies evaluating chronic effects of metformin,^34^ participants in the placebo group received a higher cumulative dose of study drug than those in the metformin group, likely due to better tolerability (Figure S1). Serum concentrations of GDF15, a metformin-sensitive biomarker and candidate mediator of its weight-reducing effect,^27^ increased significantly with metformin (Figure S2). Fourteen and eleven participants from the metformin and placebo arms, respectively, converted to diabetes and were switched to open-label metformin (p=0.50, Figure S3).

### Body weight, body composition, and metabolic indices

Both groups lost weight, but the metformin group had a greater change in weight than placebo at 24 months (−5.7 ±5.2 kg *vs*. −2.3 ±5.4 kg, p=0.0002) (Table S6). In unadjusted analysis, metformin led to change in body weight of −1.056 ±0.45 kg per year compared with placebo (p=0.0202). Weight loss with metformin was accompanied by decreases in both whole-body fat and muscle mass (Table S6). Metformin also led to significant declines in hemoglobin A1c (−0.04 ±0.3% *vs*. 0.05 ±0.3%, p=0.0229) and triglyceride concentrations (−20.1 ±43.0 mg/dL *vs*. 3.2 ±53.6 mg/dL, p=0.0096) compared with placebo.

### Frailty outcomes

For Fried phenotype, in GEE analyses piecewise linear trend with an inflection point at year 1 and autoregressive (AR)1 correlation yielded the lowest QIC. Thus, rate of change is expressed as changes in year 1 and 2. Between baseline and the 12-month follow-up, the Fried score showed an initial rate of change of 0.72 ±0.22 (p=0.0011) with metformin compared to placebo (Figure 2A), which stabilized in year 2 with a trend for a slower rate of change with metformin −0.33 ±0.17 (p=0.056). The difference in the slope of change between years 1 and 2 was significant (p=0.0026), suggesting a shift towards improvement in the Fried phenotype by year 2. When the weight loss criterion was excluded, the Fried score gradually declined in the metformin group, with no statistically significant differences in rates of change between groups: rate of change with metformin *vs*. placebo was 0.46 ±0.26 (p=0.077) in year 1 and −0.40 ±0.25 (p=0.117) in year 2 (Figure 2B). Results remained unchanged after data were censored for diabetes conversion (Figure S4). These findings suggest that the increase in the Fried phenotype score in the metformin group in year 1 was driven by significant weight loss, which stabilized in year 2.

**Figure 2.**
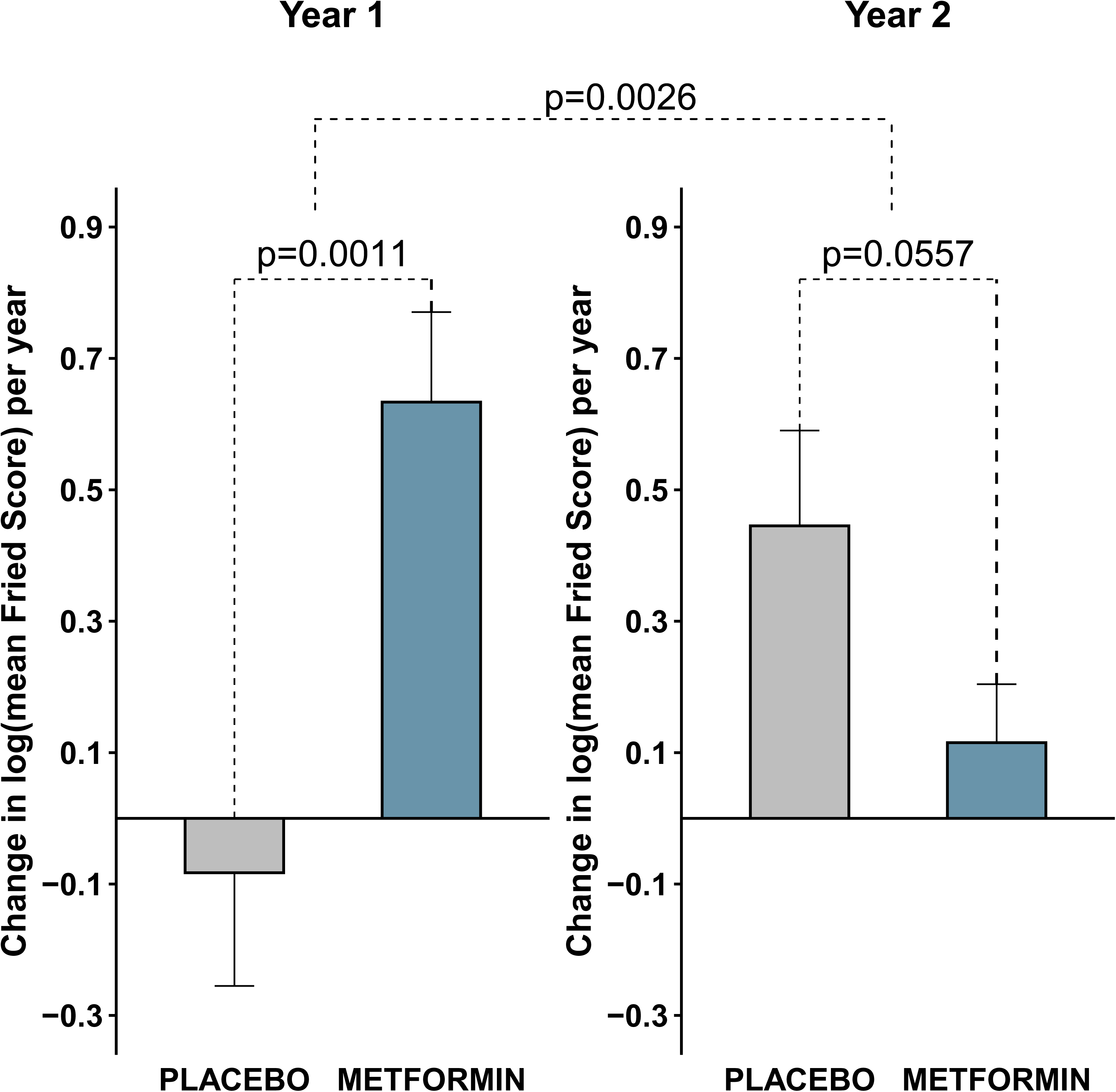
Effect of metformin on Fried frailty phenotype. Changes in Fried frailty phenotype score (0-5) by treatment group estimated using covariates-adjusted generalized estimating equations (A). Results shown as change per year for years 1 and 2 ± standard error. (B) show data for Fried phenotype without weight loss criterion.

For the frailty index, linear temporal trend and AR(1) correlation yielded the lowest QIC for frailty index. The frailty index declined in the metformin and increased in the placebo group (Figure 3A), such that metformin led to a reduction in the frailty index rate of change of −0.006 ±0.0026 per year compared to placebo in an unadjusted model (p=0.0222) (Figure 3B), which persisted after covariate adjustment (−0.005 ±0.0024, p=0.0446) and in analyses censoring data from participants after diabetes conversion (Figure S5).

**Figure 3.**
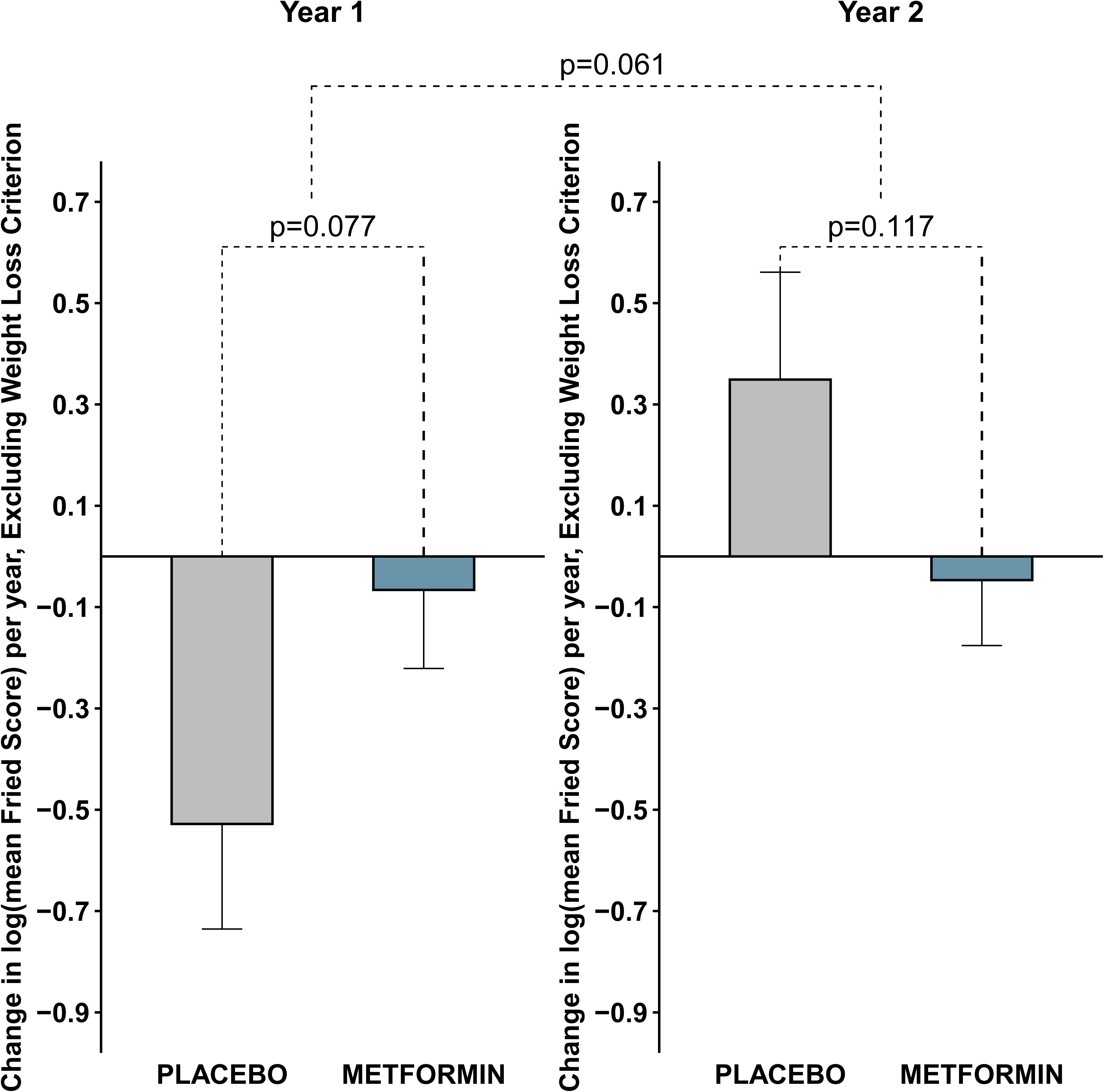
Effect of metformin vs. placebo on the deficit accumulation frailty index. Changes per year in deficit accumulation frailty index by treatment group (metformin *vs*. placebo) group, estimated using generalized estimating equations. Data shown are mean ± standard error.

Intention-to-treat analysis did not show significant changes in gait speed, grip strength, lower extremity strength, or SPPB with metformin compared to placebo (Tables S6 and S7). However, in exploratory analyses in which data were censored after diabetes conversion, significant improvement in gait speed with metformin *vs*. placebo (0.034 ±0.014, p=0.0152) was observed, which remained significant after covariate adjustment (0.033 ±0.014, p=0.0208).

### Safety and adverse events

Nearly all participants reported at least one adverse event (Table S8), the majority of which were mild and gastrointestinal in nature. Twelve participants experienced a serious adverse event and one death occurred but none were related to the study drug or intervention.

### Epigenetic clocks

All but one (PC-DNAmTL) robust clocks showed negative treatment coefficients with metformin *vs*. placebo (Figure 4). Metformin led to a significant decrease in biological age measured by PC-Hannum (−0.33 ±0.16 years, p=0.047) and PC-Horvath2 (−0.40 ±0.16 years, p=0.014) clocks *vs*. placebo. Results for the Robust Horvath Pan-Tissue Clock were only suggestive (−0.29 ±0.15 years, p=0.063). Other methylation-based clocks were not significantly affected by metformin.

**Figure 4.**
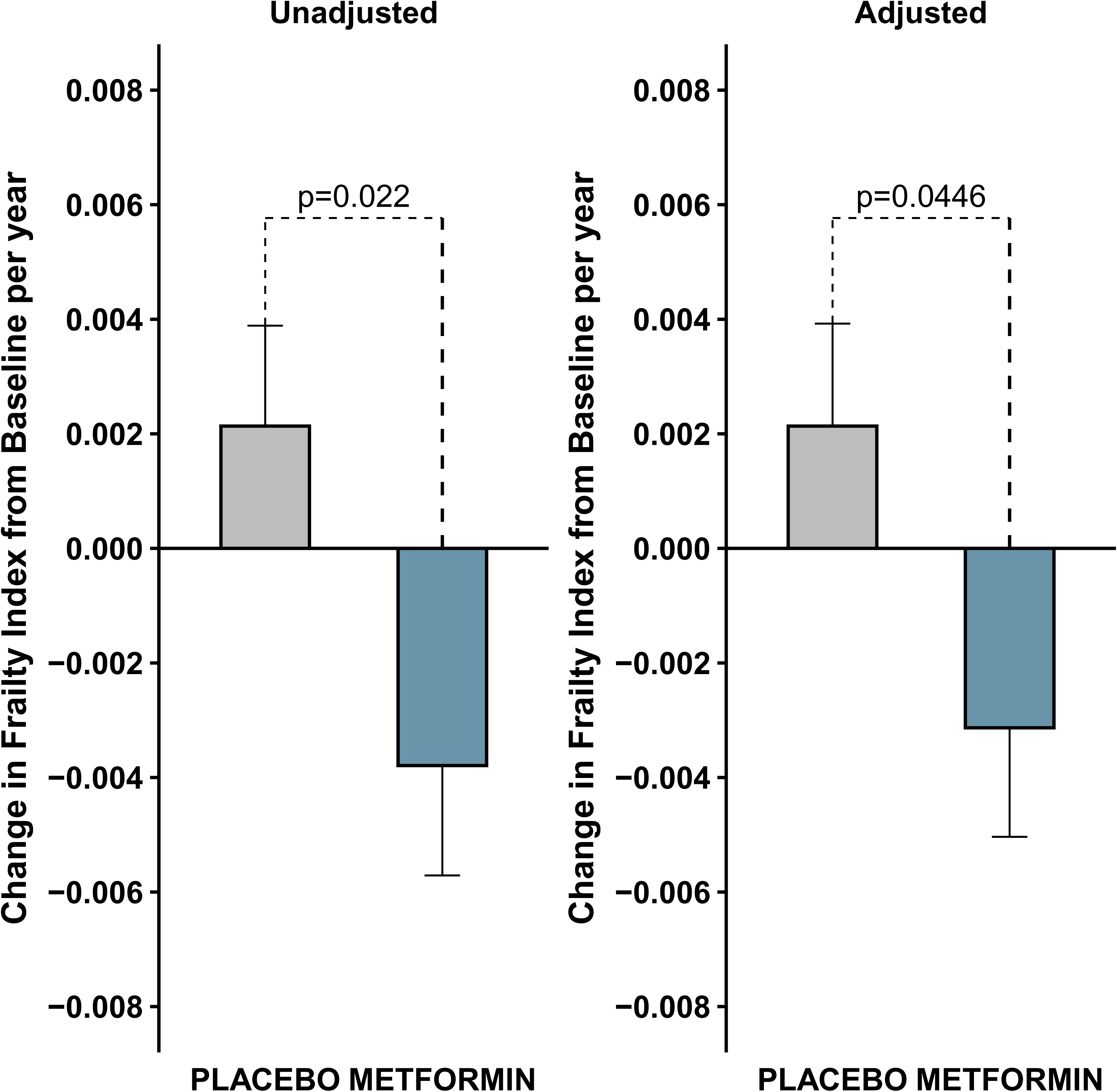
Effect of metformin on biological age. Forest plot of epigenetic clock treatment effects in blood with metformin compared to placebo.

## DISCUSSION

Metformin administration for two years reduced the deficit accumulation frailty index by 0.012 (or 0.006 per year) in older adults with glucose intolerance. To the best of our knowledge, this is the first long-term randomized trial evaluating the effect of metformin on frailty progression. This finding is clinically relevant, considering that the frailty index increases by 0.002-0.009 per year based on observational studies.^35,36^

Although several methods have been used to define frailty, the Fried phenotype^23^ and the frailty index^28^ are most commonly used and both predict poor health outcomes and mortality.^1,28,29,37–40^ The Fried phenotype conceptualizes frailty as a clinical syndrome characterized by weakness, slowness, exhaustion, unintentional weight loss, and low physical activity, and distinguishes frailty from multimorbidity and disability.^23^ In contrast, the frailty index takes a more global approach, conceptualizing frailty as the accumulation of deficits across a broad range of systems.^28^ In this study, change in the Fried phenotype was selected as the primary outcome because it was the most widely used model to quantify frailty when the trial was designed, and the majority of information about frailty prevalence and trajectory needed to inform study design was derived from the Fried phenotype.^4,24,41^ The frailty index has been increasingly recognized as a clinical marker of biological age and a useful outcome in randomized trials of various interventions, pharmacologic and lifestyle.^29,42–45^ The frailty index behaves as a continuous variable compared to the Fried^46^ and has better predictive performance in non-frail older adults.^29,37,46^ Further, the Fried phenotype considers weight loss as a deleterious phenomenon reflective of worsening frailty,^23^ while the frailty index conceptualizes major changes in body mass index - both decreases and increases - as frailty contributors. Thus, the frailty index may be more appropriate to gauge effects of interventions that change body weight on overall health.

There has been concern that interventions that cause weight loss may exacerbate sarcopenia in older adults, as both fat and lean mass are lost.^47^ Metformin resulted in decreases in muscle mass in this study; however, no significant negative effects were observed on several measures of physical function, including strength, balance, and gait speed. Further, exploratory analyses indicate an improvement in gait speed, an important indicator of lifespan and overall health.^48^

Major strengths of this study are its randomized, placebo-controlled trial design and its long duration, necessary to evaluate changes on a slowly progressing outcome such as frailty. For instance, a four month trial done in older adults with slow gait (<0.8 m/s) did not find changes with metformin,^49^ but that study might have been too short to impact frailty. Consistent with prior studies,^35^ we observed that the progression of the frailty index was relatively slow in the placebo group; it is likely that this group benefited from close medical monitoring and lifestyle counseling during the trial. Another study limitation is its modest sample size, although findings will inform the design of larger trials.

Although counterintuitive, our finding that diabetes conversion rates did not differ between the metformin and placebo groups is consistent with subgroup analyses among older adults in the Diabetes Prevention Program showing that metformin did not reduce diabetes incidence in older adults, as determined by OGTT.^50^ Albeit consistent with prior studies,^51^ the small reduction in hemoglobin A1c observed suggests metformin may reduce frailty through mechanisms other than glycemia.^7,10^ In line with this, we found metformin led to decreased biological age estimated by epigenetic clocks PC-Hannum and PC-Horvath2. Notably, robust clock measures, which use PC-based denoising to reduce technical variability in longitudinal settings, showed consistently negative treatment coefficients across all five robust clocks. Our findings are consistent with an observational study that reported metformin treatment in patients with type 2 diabetes was associated with slower epigenetic aging^52^ and vehicle-controlled studies in non-human primates indicating effects of metformin on decelerating biological age.^53^ However, next generation epigenetic clocks including GrimAge, PhenoAge, DunedinPACE, and PC-DNAmTL were not significantly affected by metformin, suggesting metformin decreases markers of biological age but does not broadly influence epigenetic predictors of mortality risk.

In conclusion, metformin slowed frailty index progression and decreased biological age in older adults. Also, metformin was safe and generally well tolerated in this population. Overall, these data support further study of metformin as a potential gerotherapeutic.

## Data Availability

All data produced in the present study are available upon reasonable request to the authors.

## Acknowledgements

We thank Beverly Orsak, RN, Alice Conde, MA, Daisy Castillo, MA, Amir Tavabi, Tiffany Cortes, MD, Rozmin Jiwani, PhD, Dean Kellogg, III, MD, Becky Powers, MD, and the research staff at Audie L. Murphy Veterans Administration Hospital and UT Health San Antonio. We also thank Dr. Joel Michalek, who assisted in preparing DSMB reports, and Drs. Jeremy Walston, George Kuchel, and Stephen Kritchevsky for their advice. We thank Karen Klein for editorial assistance. De-identified data and statistical codes will be shared upon request to the corresponding author by email.

## Data Sharing Statement

This randomized controlled trial was registered at ClinicalTrials.gov (NCT02570672) on September 8, 2015. De-identified individual participant data, the data dictionary, trial protocol, statistical analysis plan, and code used for analysis can be accessed upon request from the corresponding authors. The data will be shared under a data-sharing agreement to ensure participant confidentiality and appropriate data use. There was no patient or public involvement in the design, conduct, or reporting of this trial.

---

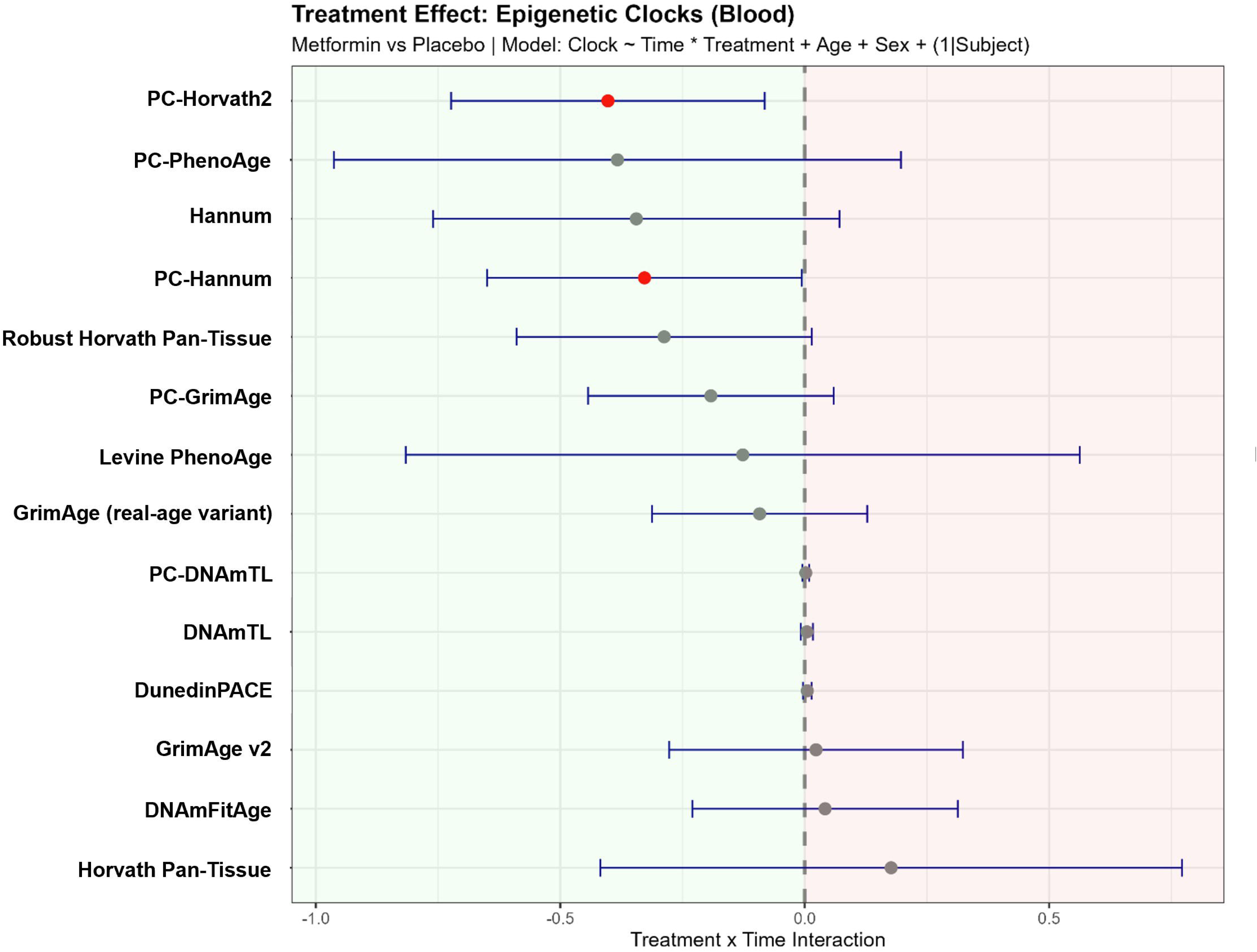

