## Supplementary Appendix for "A Randomized Clinical Trial of Metformin to Reduce Frailty in Older Adults with Glucose Intolerance"


**Table S1**. Criteria for assessing frailty by Fried phenotype definition 2

**Table S2**. Trial inclusion and exclusion criteria 4

**Table S3**. Criteria for assessing frailty by deficit accumulation frailty index 6

**Table S4**. Participant medical history of diseases and conditions at study entry 12

**Table S5**. Participant concomitant medications at study entry 13

**Figure S1**. Study drug exposure by treatment group 16

**Figure S1A**. Study drug exposure for participants randomized to metformin 16

**Figure S1B**. Study drug exposure for participants randomized to placebo 17

**Figure S1C**. Study drug exposure for participants randomized to placebo who were later switched to open-label metformin due to diabetes conversion 18

**Figure S2**. Change in GDF15 with metformin and placebo 19

**Figure S3**. Cox proportional hazard model for diabetes conversion by treatment group 20

**Table S6**. Changes in body weight, body composition, physical function, metabolic indices 21

**Figure S4**. Effect of metformin on Fried frailty with censorship for diabetes, with and without the weight loss criterion 23

**Figure S5**. Effect of metformin on frailty index, with censorship of data for diabetes 24

**Table S7**. Effect of metformin on secondary outcomes 25

**Table S8**. Summary of adverse events by treatment group 26

| **Table S1. Fried phenotype frailty criteria** | | |
|---|---|---|
| **Gait speed, 10-foot walk** | **Subgroups by height (inches)** | **Cut point for slow gait (seconds)** |
| Men | < 67.25 | ≥ 4.19 |
| | ≥ 67.5 | ≥ 3.65 |
| Women | < 61.88 | ≥ 5.31 |
| | ≥ 61.88 | ≥ 4.25 |
| **Grip strength, handheld dynamometer** | **Subgroups by body mass index (kg/m$^2$)** | **Cut point for muscle weakness (kilogram)** |
| Men | ≤ 25.1 | < 29.4 |
| | 25.2 – 27.3 | < 30.5 |
| | 27.4 – 29.8 | < 32.5 |
| | > 29.8 | <29.1 |
| Women | ≤ 25.0 | < 19.3 |
| | 25.1 – 28.1 | < 18.5 |
| | 28.2 – 31.7 | < 17.6 |
| | > 31.7 | < 17.7 |
| **Physical activity, Minnesota Leisure Time Questionnaire** | **Cut point for low physical activity (kilocalories/week)** | |
| Men | < 350 | |
| Women | < 134 | |
| **Exhaustion, Geriatric Depression Scale** | | |
| Responds "No" to question: "Do you feel full of energy?" | | |
| **Weight loss** | | |
| At baseline assessment, responds "Yes" to question: "In the last year, have you lost more than 10 pounds without trying to, that is, not by diet or exercise?" | | |

| |
|---|
| At follow-up assessments, participant experienced loss of 5% or more of body weight compared to baseline weight by direct measurement. |

| Table S2. Inclusion and exclusion criteria for trial eligibility |
|---|
| **Inclusion criteria** |
| Men and women |
| All ethnic groups |
| Age 65 and older |
| Community-dwelling |
| Pre-diabetic based on oral glucose tolerance test (OGTT) with 2-hour values of 140 – 199 mg/dL after an oral glucose load, and no diagnosis of diabetes in the past 12 months |
| Laboratory values: Hematocrit ≥ 33%, aspartate aminotransferase <twice upper limit of normal, alanine aminotransferase (ALT) < 2 X upper limit of normal, alkaline phosphatase < twice upper limit of normal, normal urinalysis (no clinically significant white blood cells, red blood cells, or bacteria), platelets ≥100,000, prothrombin < 15 seconds and prothrombin time < 40 seconds, glomerular filtration rate ≥ 45 mL/min, and urine protein ≤ 100 mg/dL by lab urinalysis |
| **Exclusion criteria** |
| Characterized as frail based on Fried phenotype criteria, defined as the presence of 3 or more of the following factors: 1) weak hand grip strength, 2) slow walking speed, 3) low physical activity, 4) unintentional weight loss of ≥ 10 lb. over the past year, 5) self-reported exhaustion |
| Resident of nursing home or long-term care facility |
| Individuals with diabetes with fasting glucose ≥ 126 mg/dL, or 2-hour glucose within diabetes range on OGTT (≥ 200 mg/dL) |
| Subjects taking drugs known to affect glucose homeostasis |
| Untreated depression or Geriatric Depression Scale score on 15-item scale >7 |
| Diagnosis of any disabling neurologic disease such as Parkinson's disease, amyotrophic lateral sclerosis, multiple sclerosis, cerebrovascular accident with residual deficits (muscle weakness or gait disorder), severe neuropathy, diagnosis of dementia or Mini-mental State Exam (MMSE) score <24, cognitive impairment due to any reason such that the person cannot provide informed consent |

| |
|---|
| History of moderate-severe heart disease (New York Heart Classification greater than grade II) or pulmonary disease (dyspnea on exertion upon climbing one flight of stairs or less; abnormal breath sounds on auscultation) |
| Poorly controlled hypertension (systolic >160 mmHg, diastolic >100 mmHg) |
| Peripheral arterial disease (history of claudication) |
| Moderate to severe valvular heart disease |

| **Table S3. Deficits assessed for the deficit accumulation frailty index** | |
|---|---|
| | **Criteria and scoring for deficit** |
| **Chronic disease or condition (medical history)** | |
| Arthritis, any type | Yes = 1; None = 0 |
| Cancer, any type | Yes = 1; None = 0 |
| Endocrine disease or condition, any type | Yes = 1; None = 0 |
| Gastrointestinal disease or condition, any type | Yes = 1; None = 0 |
| Genitourinary disease or condition, any type | Yes = 1; None = 0 |
| Head, eyes, ears, nose, throat, any disease or condition | Yes = 1; None = 0 |
| Cardiovascular or peripheral vascular disease, any type | Yes = 1; None = 0 |
| Pulmonary disease or condition, any type | Yes = 1; None = 0 |
| Hyperlipidemia | Yes = 1; None = 0 |
| Hypertension | Yes = 1; None = 0 |
| Kidney disease or condition, any type | Yes = 1; None = 0 |
| Neurological disease or condition, any type | Yes = 1; None = 0 |
| Orthopedic spine or extremity, any disease or condition | Yes = 1; None = 0 |
| Hearing difficulty | Yes = 1; None = 0 |
| Vitamin B12 deficiency | Yes = 1; None = 0 |
| **Physical examination and standardized assessments** | |
| Systolic blood pressure (mmHg) | ≥ 130 = 0.5 |
| Diastolic blood pressure (mmHg) | ≥ 80 = 0.5 |
| Body mass index (kg/$m^2$) | ≥ 30 = 1; 25 – 29.9 = 0.5; 18.5 – 24.9 = 0;<br>< 18.5 = 1 |
| Waist circumference (cm) | ≥ 102 in men = 1; ≥ 88 in women = 1 |
| Electrocardiogram | Abnormal = 1 |

| | |
|---|---|
| Time on 10-foot walk, meter/sec | ≥ 1.0 = 0; 0.8 – 0.99 = 0.33; 0.6 – 0.79 = 0.67; < 0.6 = 1 |
| Short Physical Performance Battery score | < 12 = 1 |
| ***Activities of daily living disability*** | |
| Ability to walk across a small room | Needs help = 1; Unable = 1; No help = 0 |
| Difficulty walking across a small room | A lot = 0.75; Some = 0.5; A little = 0.25; No difficulty = 0 |
| Bathing | Needs help = 1; Unable = 1; No help = 0 |
| Difficulty bathing | A lot = 0.75; Some = 0.5; A little = 0.25; No difficulty = 0 |
| Personal grooming | Needs help = 1; Unable = 1; No help = 0 |
| Difficulty with personal grooming | A lot = 0.75; Some = 0.5; A little = 0.25; No difficulty = 0 |
| Dressing | Needs help = 1; Unable = 1; No help = 0 |
| Difficulty with dressing | A lot = 0.75; Some = 0.5; A little = 0.25; No difficulty = 0 |
| Eating | Needs help = 1; Unable = 1; No help = 0 |
| Difficulty with eating | A lot = 0.75; Some = 0.5; A little = 0.25; No difficulty = 0 |
| Transferring from a bed to a chair | Needs help = 1; Unable = 1; No help = 0 |
| Difficulty transferring from a bed to a chair | A lot = 0.75; Some = 0.5; A little = 0.25; No difficulty = 0 |
| Toileting | Needs help = 1; Unable = 1; No help = 0 |
| Difficulty with toileting | A lot = 0.75; Some = 0.5; A little = 0.25; No difficulty = 0 |
| ***Instrumental activities of daily living*** | |

| | |
|---|---|
| Using the telephone | Needs help = 1; Unable = 1; No help = 0 |
| Difficulty using the telephone | A lot = 0.75; Some = 0.5; A little = 0.25; No difficulty = 0 |
| Transportation (drive or use other means of transportation) | Needs help = 1; Unable = 1; No help = 0 |
| Difficulty with transportation | A lot = 0.75; Some = 0.5; A little = 0.25; No difficulty = 0 |
| Shopping | Needs help = 1; Unable = 1; No help = 0 |
| Difficulty with shopping | A lot = 0.75; Some = 0.5; A little = 0.25; No difficulty = 0 |
| Cooking, preparing meals | Needs help = 1; Unable = 1; No help = 0 |
| Difficulty with cooking, preparing meals | A lot = 0.75; Some = 0.5; A little = 0.25; No difficulty = 0 |
| Housework | Needs help = 1; Unable = 1; No help = 0 |
| Difficulty with housework | A lot = 0.75; Some = 0.5; A little = 0.25; No difficulty = 0 |
| Taking medication | Needs help = 1; Unable = 1; No help = 0 |
| Difficulty with taking medication | A lot = 0.75; Some = 0.5; A little = 0.25; No difficulty = 0 |
| Managing finances, paying bills | Needs help = 1; Unable = 1; No help = 0 |
| Difficulty with managing finances, paying bills | A lot = 0.75; Some = 0.5; A little = 0.25; No difficulty = 0 |
| Laundry | Needs help = 1; Unable = 1; No help = 0 |
| Difficulty with laundry | A lot = 0.75; Some = 0.5; A little = 0.25; No difficulty = 0 |
| ***Short Form-12*** | |

| | |
|---|---|
| Self-reported general health | Poor = 1; Fair = 0.75; Good = 0.5; Very Good = 0.25; Excellent = 0 |
| Does your health limit you in moderate activities, such as moving a table, pushing a vacuum cleaner, bowling, or playing golf? | Yes, a lot = 1; Yes, a little = 0.5; No = 0 |
| Does your health limit you in climbing several flights of stairs? | Yes, a lot = 1; Yes, a little = 0.5; No = 0 |
| During the past 4 weeks, has your physical health resulted in you accomplishing less than you would like with your work or daily activities? | Yes, all the time = 1; Yes, most of the time = 0.75; Yes, some of the time = 0.5; Yes, a little = 0.25; No = 0 |
| During the past 4 weeks, has your physical health resulted in you being limited in the kind of work you do or other activities? | Yes, all the time = 1; Yes, most of the time = 0.75; Yes, some of the time = 0.5; Yes, a little = 0.25; No = 0 |
| During the past 4 weeks, has your emotional health resulted in you accomplishing less than you would like with your work or daily activities? | Yes, all the time = 1; Yes, most of the time = 0.75; Yes, some of the time = 0.5; Yes, a little = 0.25; No = 0 |
| During the past 4 weeks, has your emotional health resulted in you not doing work or other activities as carefully as usual? | Yes, all the time = 1; Yes, most of the time = 0.75; Yes, some of the time = 0.5; Yes, a little = 0.25; No = 0 |
| During the past 4 weeks, how much did pain interfere with your normal work (including both work outside the home and housework)? | Extremely = 1; Quite a bit = 0.75; Moderately = 0.5; A little = 0.25; Not at all = 0 |
| How much of the time have you felt calm and peaceful in the past 4 weeks? | None of the time = 1; A little of the time = 0.75; Some of the time = 0.5; A good bit of the time = 0.5; Most of the time = 0; All of the time = 0 |

| | |
|---|---|
| How much of the time did you have a lot of energy in the past 4 weeks? | None of the time = 1; A little of the time = 0.75; Some of the time = 0.5; A good bit of the time = 0.5; Most of the time = 0; All of the time = 0 |
| How much of the time have you felt downhearted and blue in the past 4 weeks? | All of the time = 1; Most of the time = 1; A good bit of the time = 0.75; Some of the time = 0.5; A little of the time = 0.25; None of the time = 0 |
| During the past 4 weeks, how much of the time has your physical health or emotional problems interfered with your social activities (like visiting with friends, relatives, *etc*.)? | All of the time = 1; Most of the time = 0.75; Some of the time = 0.5; A little of the time = 0.25; None of the time = 0 |
| ***Mini Mental State Exam*** | |
| Orientation 1 score (max score is 5) | < 5 = 1 |
| Orientation 2 score (max score is 5) | < 5 = 1 |
| Registration (max score is 3) | < 3 = 1 |
| Calculation (max score is 5) | < 5 = 1 |
| Recall (max score is 3) | < 3 = 1 |
| Language (max score is 8) | < 8 = 1 |
| Copy pentagons design (max score is 1) | < 1 = 0 |
| **Laboratory measures based on a fasting blood draw** | |
| Hemoglobin A1c (%) | > 6 = 1 |
| White blood cells ($10^9$/L) | < 4 = 1; >10 = 1 |
| Red blood cells ($10^9$L) | < 4.5 = 1; > 5.9 = 1 |
| Hemoglobin (g/dL) | < 13.5 =1; > 17.5 = 1 |
| Mean corpuscular volume (fl) | < 78 = 1; > 98 = 1 |
| Mean corpuscular hemoglobin (pg) | < 32 =1; > 36 = 1 |
| Glucose (mg/dL) | ≥ 126 = 1 |
| Urea nitrogen (mg/dL) | < 6 = 1; > 23 = 1 |

| | |
|---|---|
| Creatinine (mg/dL) | < 0.7 = 1; > 1.2 = 1 |
| Sodium (mmol/L) | < 133 = 1; > 145 =1 |
| Calcium (mg/dL) | < 8.6 = 1; > 10.3 = 1 |
| Protein, total (g/dL) | < 6.4 = 1; > 8.9 = 1 |
| Albumin (g/dL) | < 3.5 =1; > 5.7 = 1 |
| Alkaline phosphatase (IU/L) | < 34 = 1; > 104 = 1 |
| Total bilirubin (mg/dL) | < 0.3 =1; > 1.0 = 1 |
| Cholesterol (mg/dL) | > 200 = 1 |
| Triglycerides (mg/dL) | > 200 = 1 |
| HDL (mg/dL) | < 23 = 1 |
| LDL (mg/dL) | > 193 = 1 |
| Prothrombin (sec) | < 10.1 = 1; > 13.0 = 1 |
| International Normalized Ratio | < 0.8 = 1; > 1.2 = 1 |
| Prothrombin time (sec) | < 25.1 = 1; > 36.5 = 1 |
| Urinalysis, red blood cells | Abnormal = 1; Normal = 0 |
| Vitamin B12 level (pg/mL) | < 180 = 1 |
| **For follow-up assessments, the items below were added** | |
| Hematocrit (%) | < 41 = 1; > 53 = 1 |
| Platelets | < 150 = 1; > 400 = 1 |
| Potassium (mmol/L) | < 3.5 = 1; > 5.0 = 1 |
| Aspartate transferase (IU/L) | < 13 = 1; > 39 = 1 |
| Alanine aminotransferase (IU/L) | < 7 = 1; > 52 = 1 |
| Geriatric Depression Scale, score | > 7 = 1 |
| Urinalysis | Abnormal = 1 |
| Urine protein | Abnormal = 1 |
| Oral glucose tolerance test, 2-hour glucose | > 200 = 1 |

| Table S4. Medical history of chronic diseases and conditions of participants at baseline by self-report | | | |
|---|---|---|---|
| | **Metformin**<br>N= 70 | **Placebo**<br>N= 71 | **Overall**<br>N= 141 |
| **Characteristic** | *N (%)* | | |
| Hypertension | 33 (47) | 42 (59) | 75 (53) |
| Hypercholesterolemia, hyperlipidemia | 44 (63) | 51 (72) | 95 (67) |
| Arthritis, any type | 28 (40) | 39 (55) | 67 (48) |
| History of cancer | 15 (21) | 13 (18) | 28 (20) |
| Endocrine disorders, not diabetes | 32 (46) | 26 (37) | 58 (41) |
| Gastrointestinal | 37 (53) | 45 (63) | 82 (58) |
| Genitourinary | 26 (37) | 25 (35) | 51 (36) |
| Cardiovascular diseases (central, not peripheral) | 36 (51) | 40 (56) | 76 (54) |
| Peripheral vascular disease | 14 (20) | 19 (27) | 33 (23) |
| Pulmonary | 9 (13) | 21 (30) | 30 (21) |
| Neurologic | 17 (24) | 20 (28) | 37 (26) |
| Sleep disorders | 18 (26) | 13 (18) | 31 (22) |
| Orthopedic, spine | 13 (19) | 15 (21) | 28 (20) |
| Orthopedic, extremity | 36 (51) | 40 (56) | 76 (54) |
| Chronic pain | 29 (41) | 19 (27) | 48 (34) |
| Mental health condition | 19 (27) | 19 (27) | 38 (27) |

| Table S5. Baseline concomitant medications at study entry* | | |
|---|---|---|
| | **Metformin**<br><br>N = 70 | **Placebo**<br><br>N = 71 |
| **Drug category** | *N (%)* | |
| **Analgesic** | 44 (62.9) | 47 (66.2) |
| Nonsteroidal anti-inflammatory drug | 32 | 30 |
| Nonopioid | 27 | 18 |
| Topical | 2 | 3 |
| Topical, local anesthetic | 2 | 1 |
| Nonsteroidal anti-inflammatory drug, COX-2 selective | 1 | 1 |
| Nonsteroidal anti-inflammatory drug, topical | 1 | 1 |
| Opioid | 1 | 3 |
| Salicylate | 1 | 0 |
| Urinary | 1 | 0 |
| Nonopioid / opioid | 0 | 1 |
| **Antihypertensive** | 34 (48.6) | 47 (66.2) |
| Angiotensin II receptor blocker | 13 | 14 |
| Calcium channel blocker | 12 | 13 |
| Beta blocker | 8 | 20 |
| Angiotensin-converting enzyme (ACE) inhibitor | 6 | 14 |
| Diuretic, thiazide | 4 | 11 |
| Angiotensin II receptor blocker/Diuretic, thiazide | 3 | 5 |
| Diuretic, loop | 3 | 1 |
| Angiotensin-converting enzyme (ACE) inhibitor/Diuretic, thiazide | 2 | 2 |
| Beta blocker with alpha blocking activity | 2 | 2 |

| | | |
|---|---|---|
| Beta blocker/Diuretic, thiazide | 1 | 0 |
| Diuretic, thiazide/Diuretic, potassium sparing | 1 | 0 |
| Vasodilator | 0 | 1 |
| **Antilipemic agent** | 42 (60) | 52 (73.2) |
| HMG-CoA reductase inhibitor | 36 | 46 |
| Omega-3 fatty acids | 24 | 17 |
| 2-Azetidinone | 1 | 2 |
| PCSK9 inhibitor | 0 | 1 |
| Vitamin, water soluble | 0 | 1 |
| **Antiplatelet agent** | 23 (32.9) | 32 (45.1) |
| Salicylate | 23 | 30 |
| P2Y12 Antagonist | 3 | 3 |
| **Dietary supplement** | 37 (52.9) | 33 (46.5) |
| Ubiquinol | 13 | 7 |
| **Electrolyte supplement** | 27 (38.6) | 26 (36.6) |
| Calcium salt | 17 | 14 |
| Ophthalmic | 3 | 4 |
| Intranasal | 2 | 0 |
| Topical | 1 | 0 |
| **Histamine H1 antagonist** | 27 (38.6) | 31 (43.7) |
| Intranasal | 3 | 4 |
| Ophthalmic | 2 | 0 |
| **Thyroid product** | 17 (24.3) | 15 (21.1) |
| **Vaccine** | 20 (28.6) | 18 (25.4) |
| **Vitamin** | 47 (67.1) | 40 (56.3) |
| Vitamin, water soluble | 32 | 22 |

| | | |
|---|---|---|
| Multivitamin | 28 | 26 |
| Ophthalmic | 3 | 2 |
| Vitamin, fat soluble | 1 | 3 |
| **Vitamin D analog** | 28 (40) | 32 (45.1) |

* Drug categories for medications used in ≥20% of participants.

**Figure S1**. Average daily dosage for participants who took at least one dose of study drug. Data are shown for participants randomized to metformin (Figure S1A), placebo (Figure S1B), and those randomized to placebo but switched to open-label metformin due to diabetes conversion (Figure S1C).

**Figure S1A**.

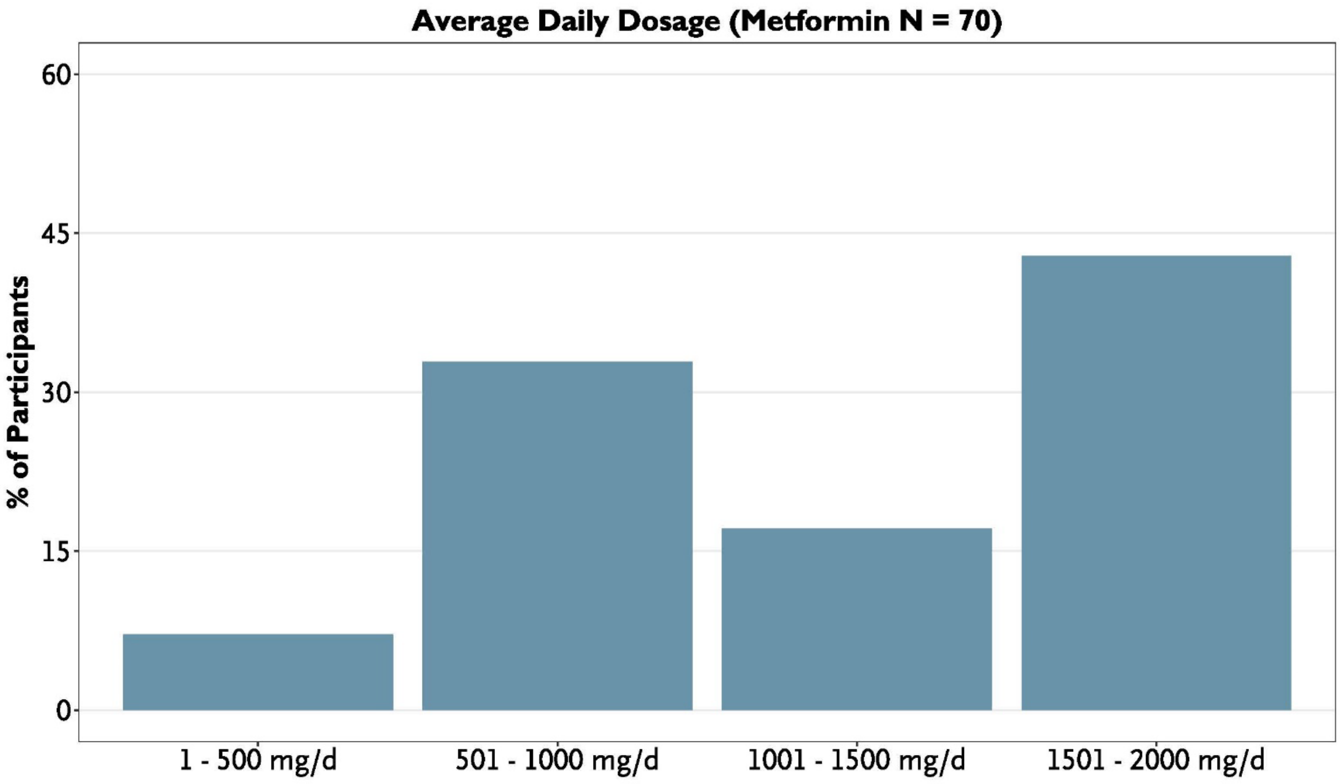

**Figure S1B**.

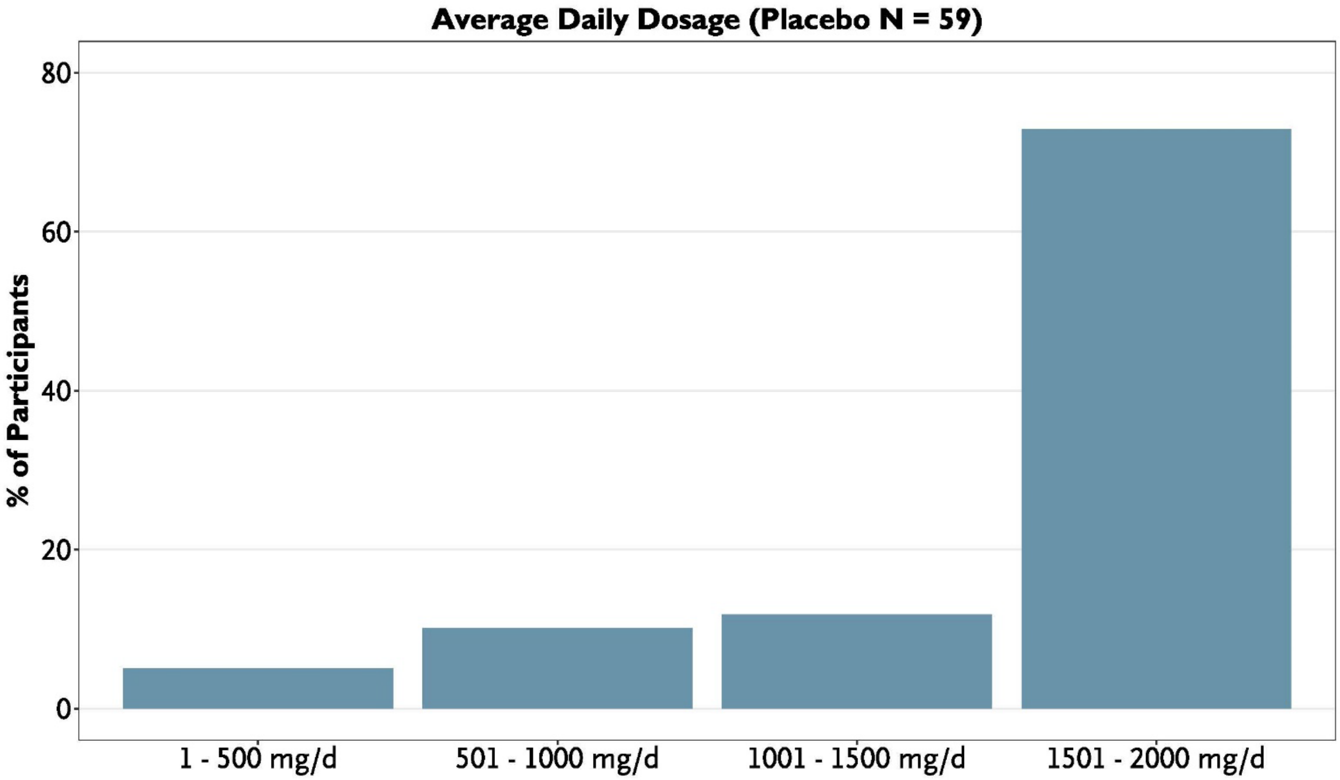

Average Daily Dosage (Placebo N = 59)
80
60
40
20
0
% of Participants
1 - 500 mg/d
501 - 1000 mg/d
1001 - 1500 mg/d
1501 - 2000 mg/d

**Figure S1C**.

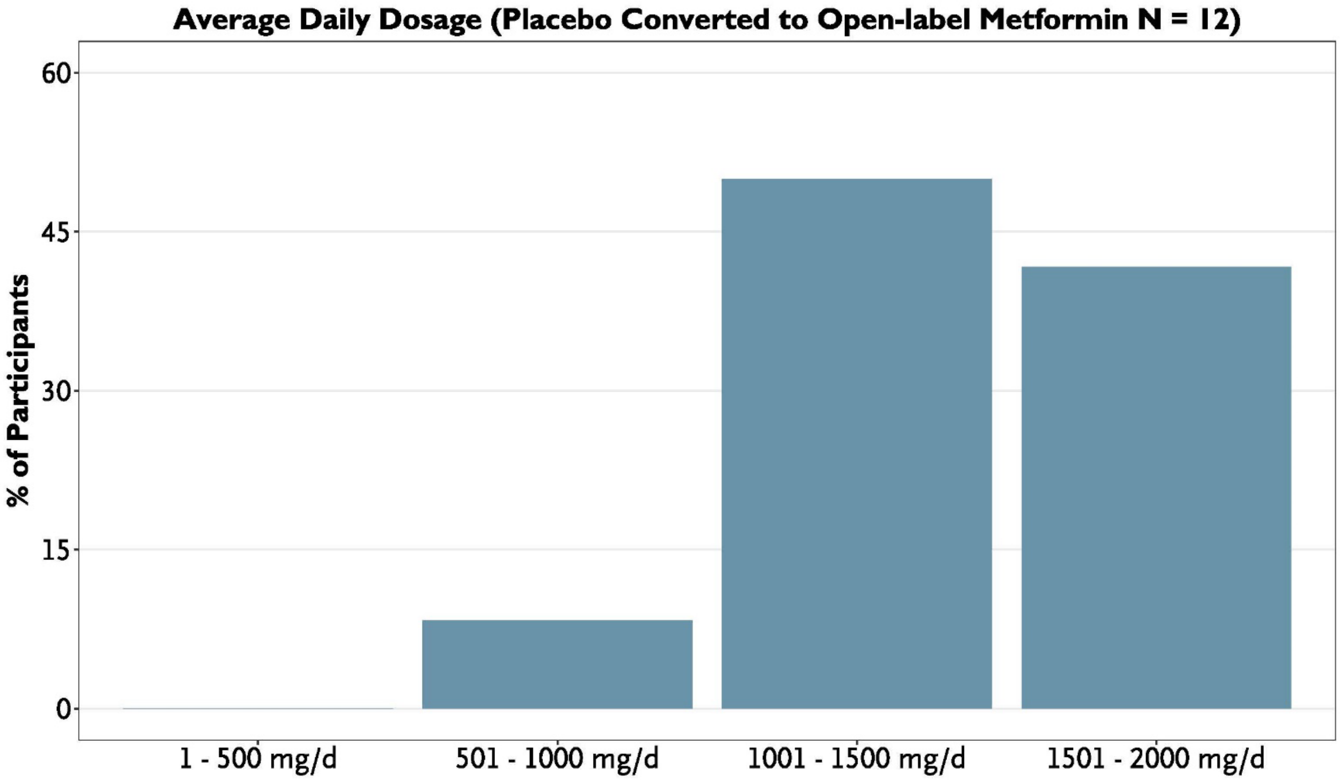
Average Daily Dosage (Placebo Converted to Open-label Metformin N = 12)
% of Participants
60
45
30
15
0
1 - 500 mg/d
501 - 1000 mg/d
1001 - 1500 mg/d
1501 - 2000 mg/d

**Figure S2.** Change in serum GDF15 concentrations. Data are means ± standard deviation.

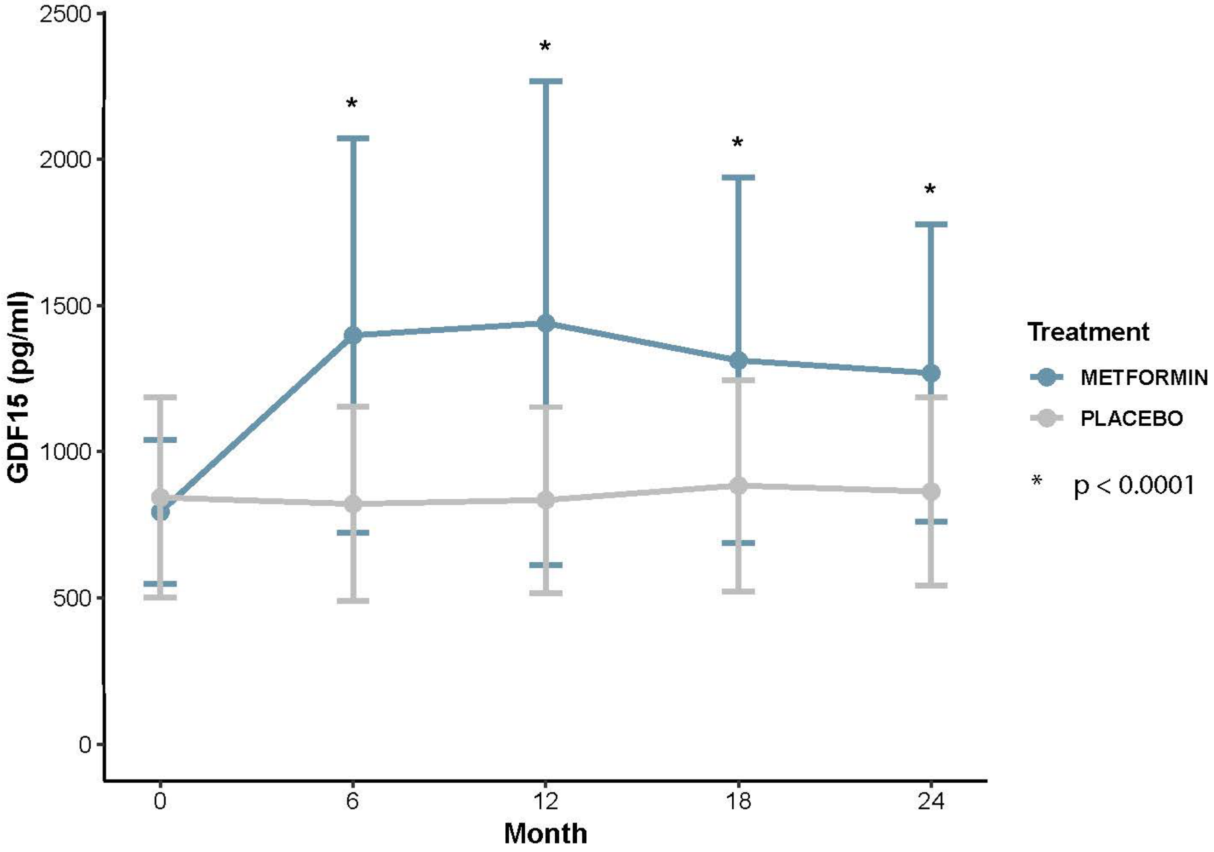

**Figure S3**. Cox proportional hazards model for diabetes conversion by treatment group (metformin *vs*. placebo) during study follow-up.

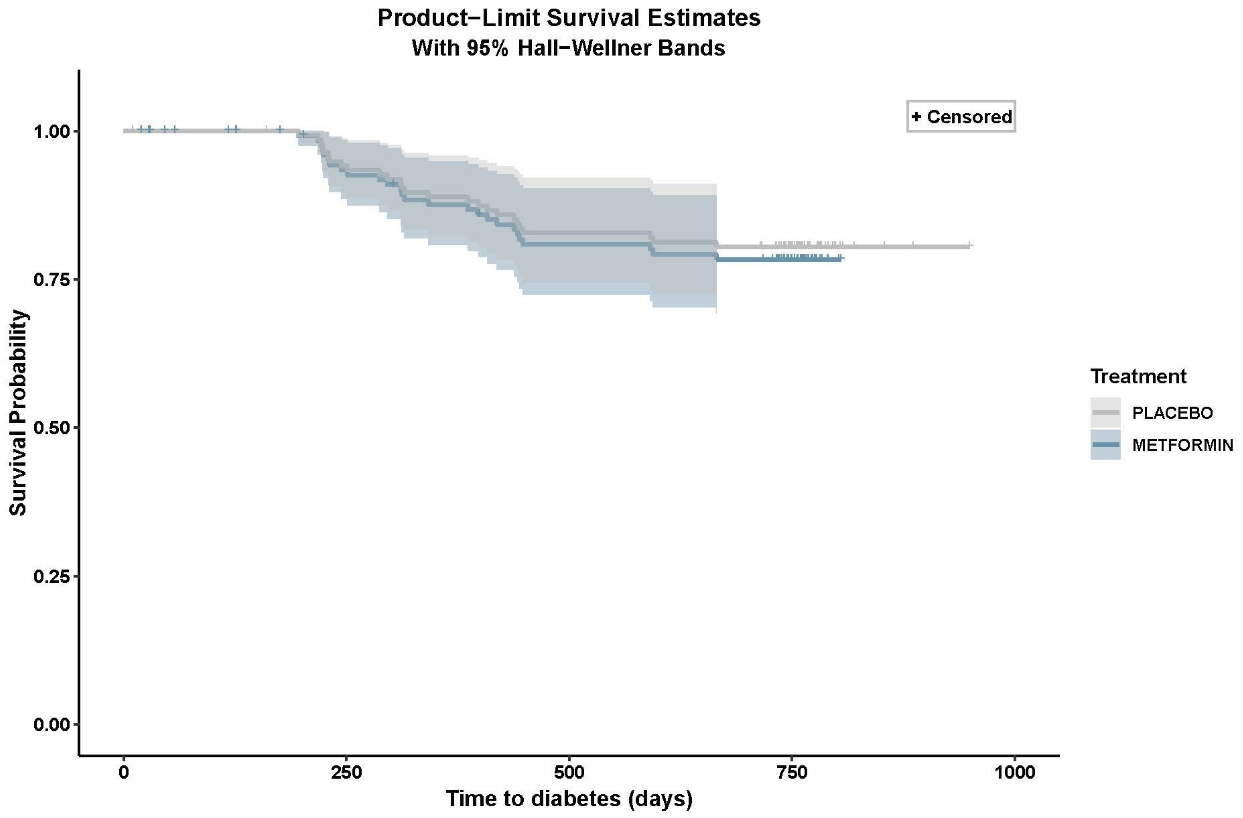

| **Table S6. Changes in body weight, blood pressure, metabolic indices and physical function** | | | | | | |
|---|---|---|---|---|---|---|
| | **Baseline to 12 months** | | | **Baseline to 24 months** | | |
| | **Metformin** | **Placebo** | **P*** | **Metformin** | **Placebo** | **P*** |
| | Mean Change (SD) | Mean Change (SD) | | Mean Change (SD) | Mean Change (SD) | |
| Body weight, kg | -4.78 (4.52) | -1.64 (4.52) | 7.321E-05 | -5.70 (5.19) | -2.26 (5.37) | 0.0002 |
| Body mass index, kg/m$^2$ | -1.68 (1.57) | -0.56 (1.64) | 0.0001 | -2.02 (1.89) | -0.82 (1.93) | 0.0004 |
| Systolic blood pressure, mmHg | -5.26 (16.32) | -8.88 (16.10) | 0.3298 | -0.90 (16.57) | -7.17 (18.69) | 0.0596 |
| Diastolic blood pressure, mmHg | -2.25 (9.32) | -2.77 (10.31) | 0.7716 | -0.22 (8.76) | -2.11 (9.59) | 0.3271 |
| Fasting plasma glucose, mg/dL | -3.33 (9.96) | -1.65 (16.05) | 0.1000 | -2.78 (9.96) | -0.75 (10.74) | 0.0961 |
| 2-hour glucose on OGTT, mg/dL | -2.98 (40.30) | -9.81 (31.27) | 0.4902 | -0.11 (36.30) | -8.4 (39.39) | 0.1343 |
| Hemoglobin A1c, % | -0.042 (0.22) | 0.0044 (0.25) | 0.1088 | -0.04 (0.25) | 0.05 (0.31) | 0.0229 |
| Total cholesterol, mg/dL | -12.95 (33.32) | -6.5 (25.38) | 0.1470 | -13.07 (30.00) | -9.43 (31.68) | 0.6070 |
| Triglycerides, mg/dL | -16.23 (46.54) | 5.46 (38.10) | 0.0293 | -20.07 (42.68) | 3.17 (53.60) | 0.0096 |
| Total fat mass, g | -2205.94 (4685.03) | -437.23 (4336.36) | 0.0505 | -3023.74 (5191.15) | -948.25 (4492.93) | 0.0227 |
| Total muscle mass, g | -2771.74 (7354.58) | -1300.52 (4411.59) | 0.0978 | -2125.31 (5152.68) | -1381.48 (4240.02) | 0.0496 |
| Visceral fat mass, g | -13.95 | -5.62 | 0.6341 | -45.55 | -30.83 | 0.6128 |

| | | | | | | |
|---|---|---|---|---|---|---|
| | (215.85) | (203.12) | | (213.71) | (193.30) | |
| Gait speed, meters/second | 0.00308 (0.1816) | -0.03998 (0.16116) | 0.1324 | -0.01623 (0.1632) | -0.0665 (0.1823) | 0.1464 |
| 6-minute walk test, meters | 24.5641 (43.5711) | 5.4298 (48.1947) | 0.03259 | 17.1621 (48.1748) | 2.4162 (37.7506) | 0.2158 |
| Grip strength, kg | -0.6835 (3.9047) | -0.2967 (5.8042) | 0.2963 | -1.3472 (3.6862) | 0.314 (4.3018) | 0.06248 |
| Lower extremity strength, Nm | -3.2657 (19.6897) | -3.0744 (23.2242) | 0.8881 | -4.7912 (18.4349) | 3.2789 (26.4662) | 0.2004 |

Values are unadjusted changes. *P-value from Wilcoxon rank-sum test for treatment group comparisons.

**Figure S4. Effect of metformin *vs*. placebo on Fried frailty phenotype with censorship of observations after diabetes conversion**. Yearly change in Fried frailty phenotype score by treatment group (A). Yearly change in Fried frailty score by treatment group without weight loss criterion (B). Data shown represent estimated yearly change ± standard error.

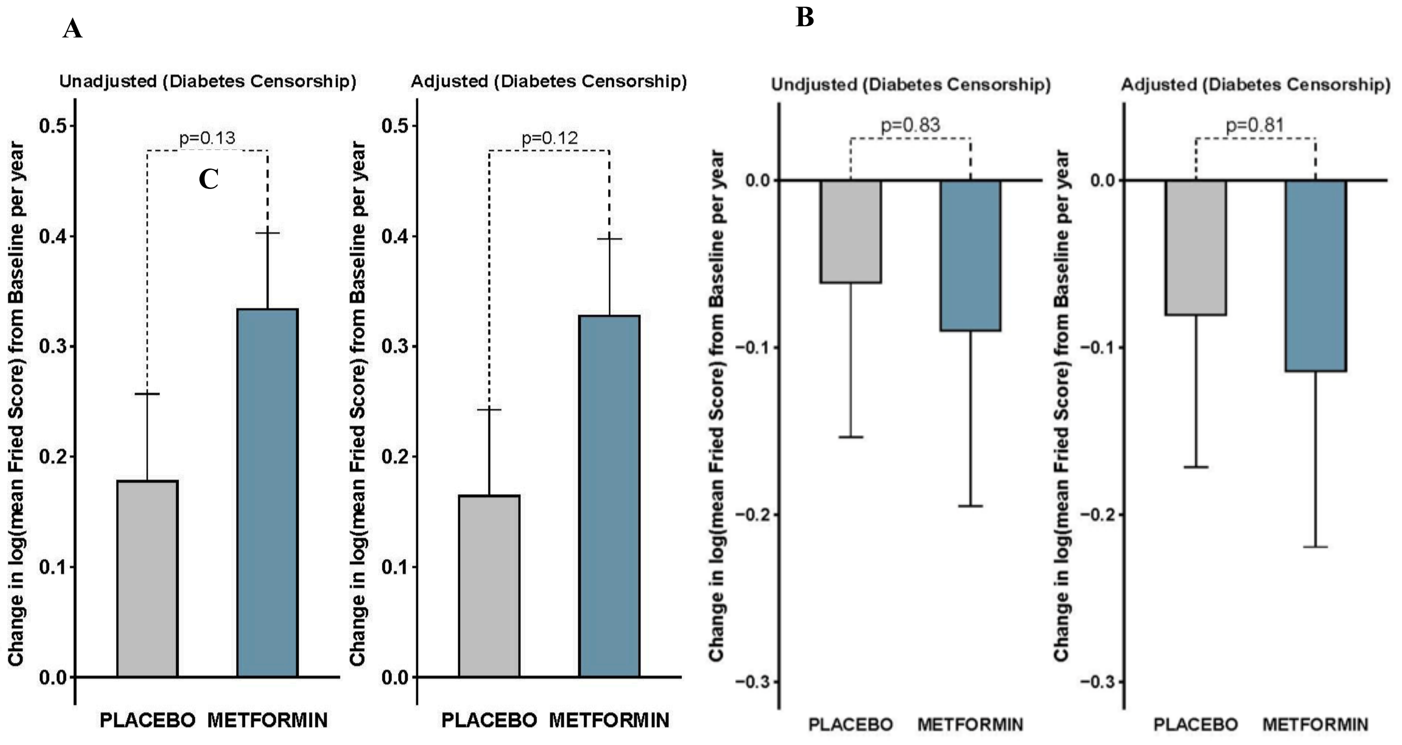

**Figure S5. Effect of metformin *vs*. placebo on deficit accumulation frailty index, with censorship of observations after diabetes conversion**. Yearly change in deficit accumulation frailty index by treatment group based on generalized estimating equations model results. Data shown represent estimated yearly change ± standard error.

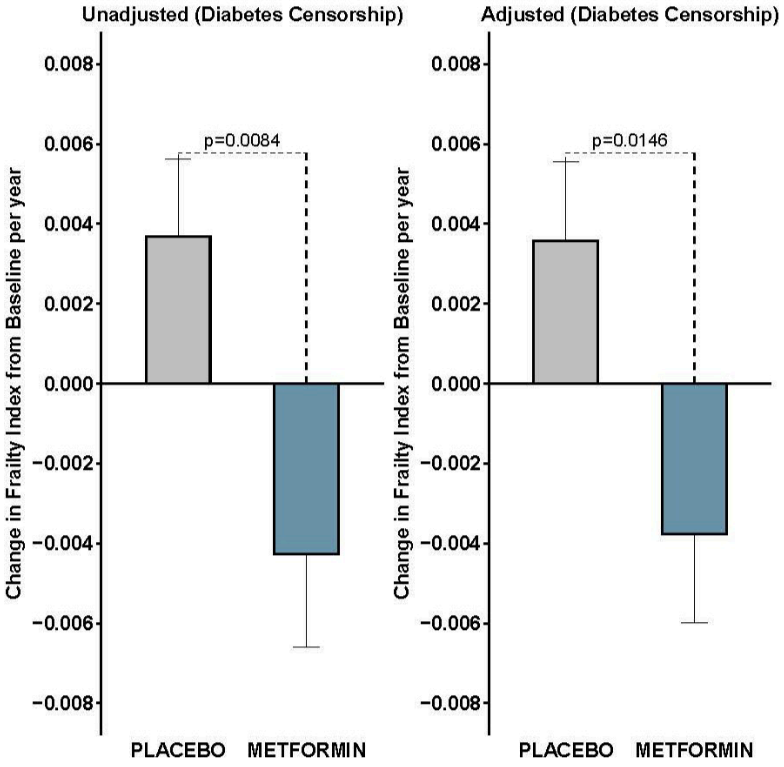

| **Table S7. Changes in frailty index and physical function outcomes per year with metformin *vs*. placebo based on generalized estimating equations (unadjusted values).** | | | | | | |
|---|---|---|---|---|---|---|
| | **Intention to Treat** | | | **Censored for Diabetes Conversion** | | |
| | **Estimate (SE)** | **CI** | **P-value** | **Estimate (SE)** | **CI** | **P-value** |
| **Frailty index** | -0.0059 (0.0026) | -0.0109, -0.0008 | 0.022 | -0.0079 (0.0030) | -0.0138, -0.0019 | 0.0093 |
| **Gait speed, meter/second** | 0.024 (0.013) | -0.0008, 0.049 | 0.058 | 0.034 (0.014) | 0.0064, 0.061 | 0.0152 |
| **Grip strength, kg** | -0.49 (0.39) | -1.26, 0.28 | 0.22 | -0.27 (0.46) | -1.17, 0.63 | 0.5508 |
| **Lower extremity strength, Nm** | 6.85 (8.55) | -9.91, 23.60 | 0.4233 | 9.91 (9.55) | -8.80, 28.62 | 0.2993 |
| **Short Physical Performance Battery** Unadjusted | 0.0397 (0.13) | -0.22, 0.295 | 0.7607 | -0.0268 (0.146) | -0.3129, 0.2593 | 0.8544 |

| Table S8. Summary of adverse events | | | |
|---|---|---|---|
| | **Metformin** | **Placebo** | |
| | N = 70 | N = 71 | |
| | | **Placebo** | **Placebo to Open-label Metformin** |
| | | N = 59 | N = 12 |
| | *Number of participants (%)* | | |
| **Any adverse event** | 70 (100) | 59 (100) | 11 (91.7) |
| **Adverse events by system organ class*** | | | |
| Blood and lymphatic system disorders | 2 (2.9) | 1 (1.7) | 2 (16.7) |
| Cardiac disorders | 23 (32.9) | 21 (35.6) | 6 (50) |
| Congenital, familial and genetic disorders | 2 (2.9) | 1 (1.7) | 0 (0) |
| Ear and labyrinth disorders | 5 (7.1) | 6 (10.2) | 1 (8.3) |
| Endocrine disorders | 2 (2.9) | 1 (1.7) | 0 (0) |
| Eye disorders | 10 (14.3) | 11 (18.6) | 5 (41.7) |
| Gastrointestinal disorders | 63 (90.0) | 40 (67.8) | 10 (83.3) |
| General disorders and administration site conditions | 41 (58.6) | 31 (52.5) | 8 (66.7) |
| Hepatobiliary disorders | 4 (5.7) | 2 (3.4) | 0 (0) |
| Immune system disorders | 7 (10.0) | 11 (18.6) | 1 (8.3) |
| Infections and infestations | 33 (47.1) | 38 (64.4) | 5 (41.7) |
| Injury, poisoning and procedural complications | 40 (57.1) | 40 (67.8) | 7 (58.3) |
| Investigations | 34 (48.6) | 29 (49.2) | 6 (50) |
| Metabolism and nutrition disorders | 7 (10.0) | 7 (11.9) | 4 (33.3) |

| | | | |
|---|---|---|---|
| Musculoskeletal and connective tissue disorders | 50 (71.4) | 52 (88.1) | 8 (66.7) |
| Neoplasms benign, malignant and unspecified (including cysts and polyps) | 8 (11.4) | 9 (15.3) | 4 (33.3) |
| Nervous system disorders | 45 (64.3) | 28 (47.5) | 7 (58.3) |
| Psychiatric disorders | 13 (18.6) | 10 (16.9) | 3 (25) |
| Renal and urinary disorders | 11 (15.7) | 12 (20.3) | 1 (8.3) |
| Reproductive system and breast disorders | 3 (4.3) | 3 (5.1) | 0 (0) |
| Respiratory, thoracic and mediastinal disorders | 21 (30.0) | 32 (54.2) | 4 (33.3) |
| Skin and subcutaneous tissue disorders | 18 (25.7) | 25 (42.4) | 2 (16.7) |
| Surgical and medical procedures | 27 (38.6) | 20 (33.9) | 2 (16.7) |
| Vascular disorders | 8 (11.4) | 18 (30.5) | 0 (0) |
| **Most frequent adverse events by preferred term**† | **Metformin** | **Placebo** | **Placebo to Open-label Metformin** |
| | *Number of participants (%)* | | |
| Diarrhea | 50 (71.4) | 24 (40.7) | 9 (75.0) |
| Arthralgia | 22 (31.4) | 21 (35.6) | 3 (25.0) |
| Nausea | 21 (30.0) | 8 (13.6) | 4 (33.3) |
| Flatulence | 15 (21.4) | 5 (8.5) | 2 (16.7) |
| Edema peripheral | 15 (21.4) | 11 (18.6) | 2 (16.7) |
| Pain in extremity | 15 (21.4) | 13 (22) | 2 (16.7) |
| Contusion | 14 (20.0) | 14 (23.7) | 3 (25) |
| Headache | 14 (20.0) | 8 (13.6) | 2 (16.7) |

| | | | |
|---|---|---|---|
| Dizziness | 13 (18.6) | 4 (6.8) | 5 (41.7) |
| Constipation | 8 (11.4) | 9 (15.3) | 4 (33.3) |
| Cough | 8 (11.4) | 12 (20.3) | 1 (8.3) |
| Back pain | 7 (10.0) | 11 (18.6) | 3 (25) |
| Balance disorder | 5 (7.1) | 6 (10.2) | 3 (25) |
| Decreased appetite | 4 (5.7) | 0 (0) | 3 (25) |
| **Intensity of adverse events instances**‡ | **Metformin** | **Placebo** | **Placebo to Open-label Metformin** |
| | Instance frequency = 1,214 | Instance frequency = 1,009 | Instance frequency = 233 |
| Mild | 736 (60.6) | 605 (60) | 151 (64.8) |
| Moderate | 421 (34.7) | 361 (35.8) | 69 (29.6) |
| Severe | 57 (4.7) | 43 (4.3) | 13 (5.6) |
| | **Metformin** | **Placebo** | **Placebo to Open-label Metformin** |
| | N = 70 | N = 59 | N = 12 |
| | *Number of participants (%)* | | |
| **Serious adverse events** § | 8 (11.4) | 3 (5.1) | 1 (8.3) |
| Serious adverse event considered by the investigator to be related to metformin or placebo | 0 | 0 | 0 |
| Serious adverse event resulting in hospitalization | 8 | 3 | 1 |
| Serious adverse event resulting in admission to a rehabilitation facility | 0 | 0 | 0 |

| | | | |
|---|---|---|---|
| Serious adverse event resulting in admission to a long-term nursing facility | 0 | 0 | 0 |

* Chi-squared test result for difference in participants experiencing adverse events by group (metformin, placebo, placebo to open-label metformin): X-squared = 49.33, degrees of freedom = 46, p = 0.3414

† Includes adverse events by preferred term with incidence of 20% or higher. Chi-squared test result for differences in participants experiencing adverse events by group (metformin, placebo, placebo to open-label metformin): X-squared = 36.784, degrees of freedom = 26, p = 0.07817

‡ Chi-squared test result for differences in adverse event instance intensity by group (metformin, placebo, placebo to open-label metformin): X-squared = 3.575, degrees of freedom = 4, p = 0.4666

§ Chi-squared test result for differences among participants experiencing serious adverse events by group (metformin, placebo, placebo to open-label metformin): X-squared = 1.5144, degrees of freedom = 2, p = 0.469
